# Cell-Free DNA Genomic and Fragmentomic Features for Early Outcome Prediction in Large B-Cell Lymphoma

**DOI:** 10.64898/2026.05.29.26353426

**Authors:** Steven Wang, Parisa Mapar, Norbert Moldovan, Ymke van der Pol, Aram Safrastyan, Erik van Werkhoven, Normastuti Adhini Tantyo, Bart Snieder, André F. Do Brito Valente, A. Vera de Jonge, Avinash Dinmohamed, Esther E.E. Drees, Margaretha G.M. Roemer, Bauke Ylstra, Clara P.W. Klerk, Leonie Strobbe, Yorick Sandberg, Rinske S. Boersma, Harry Koene, Hans Pruijt, Koen de Heer, Rozemarijn van Rijn, Yavuz M. Bilgin, Eva de Jongh, Marcel Nijland, Marjolein van der Poel, Ad Koster, Laurens Nieuwenhuizen, Rob Fijnheer, Aart Beeker, Rogier Mous, Vibeke K.J. Vergote, Joost S.P. Vermaat, D. Michiel Pegtel, Martine E.D. Chamuleau, Florent Mouliere

**Affiliations:** Amsterdam UMC Location Vrije Universiteit Amsterdam, Hematology, Amsterdam, The Netherlands; Amsterdam UMC Location Vrije Universiteit Amsterdam, Pathology, Amsterdam, The Netherlands; Cancer Center Amsterdam, Imaging and Biomarkers, Amsterdam, The Netherlands; Institute for Molecular Medicine Finland (FIMM), HiLIFE, University of Helsinki, Finland; Cancer Research UK National Biomarker Centre, University of Manchester, Manchester, United Kingdom; Hemato-Oncology Foundation for Adults in The Netherlands (HOVON), The Netherlands; Netherlands Comprehensive Cancer Organization (IKNL), The Netherlands; Dijklander Hospital, Hoorn, The Netherlands; Gelre Ziekenhuis, Zutphen, The Netherlands; Maasstad Ziekenhuis, Rotterdam, The Netherlands; Amphia Ziekenhuis, Breda, The Netherlands; St. Antonius Ziekenhuis, Nieuwegein, The Netherlands; Jeroen Bosch Ziekenhuis, ’s-Hertogenbosch, The Netherlands; Flevoziekenhuis, Almere, The Netherlands; Medical Center Leeuwarden, Leeuwarden, The Netherlands; Adrz, Goes, The Netherlands; Albert Schweitzer Ziekenhuis, Dordrecht, The Netherlands; University Medical Center Groningen, Groningen, The Netherlands; Maastricht University Medical Center, Maastricht, The Netherlands; VieCuri Medical Center, Venlo, The Netherlands; Maxima Medical Center, Eindhoven, The Netherlands; Meander Medical Center, Amersfoort, The Netherlands; Spaarne Gasthuis, Hoofddorp, The Netherlands; University Medical Center Utrecht, Utrecht, The Netherlands; University Hospitals Leuven, Leuven, Belgium; Leiden University Medical Center, Leiden, The Netherlands

**Keywords:** cell-free DNA, fragmentomics, large B-cell lymphoma, liquid biopsy, shallow whole-genome sequencing, risk stratification, multimodal

## Abstract

Curative-intent immunochemotherapy fails in ∼30% of patients with large B-cell lymphoma (LBCL), yet no validated molecular tool enables early identification of high-risk individuals to guide treatment intensification. Using shallow whole genome sequencing (sWGS) of plasma cell-free DNA from 190 LBCL patients, we developed and validated the ACT score (Aberrations, fragment Composition, Terminal motifs), a composite classifier integrating genomic and fragmentomic features from a single post-cycle-1 sample. ACT-positive patients had worse 2-year outcomes versus ACT-negative patients: time-to-progression 29% vs. 83% (HR 4.4, 95% CI 1.9–10.0; *P* = 1.5 × 10^-4^) and overall survival 47% vs. 93% (HR 8.7, 95% CI 3.0–25.4; *P* = 1.8 × 10^-6^). ACT score was independently prognostic of the International Prognostic Index, and their combination identified the highest-risk patients. Unlike mutation-based approaches, this assay requires neither tumor tissue, germline control nor a baseline plasma sample. Built on open-source tools and sWGS, the ACT score offers a feasible scalable strategy for early risk stratification in aggressive LBCL.

**Highlights:**

- Early identification of DLBCL patients likely to fail first-line treatment remains challenging
- ACT score (Aberrations, Composition of fragments, Terminal motif analyses) integrates cell-free DNA genomic and fragmentomic features from a single plasma sample at cycle 2 day 1
- Tumor-naive, open-source, WGS approach without needing tissue or germline control
- ACT score in combination with International Prognostic Index enables early risk stratification

## Introduction

Despite advances in first-line immunochemotherapy, up to 30% of patients with large B-cell lymphoma (LBCL) develop relapsed or refractory (R/R) disease, where outcomes remain poor.^1–3^ Clinical trials using classic risk stratification tools including the International Prognostic Index (IPI), cell-of-origin (COO) classification, and interim PET-CT (iPET) have had limited success in improving outcome.^4–10^ At the same time, novel salvage therapies including bispecific antibodies and CAR-T cell therapy are reshaping outcomes in R/R disease,^11–13^ underscoring the urgency of biomarkers that can accurately identify high-risk individuals who are most likely to benefit from early therapy escalation.^14^

Cell-free DNA (cfDNA) has emerged as a powerful liquid biopsy tool in solid tumors,^15,16^ and its role in aggressive lymphomas is an active field of intense investigation. Tumor-informed sequencing strategies enable sensitive measurable residual disease (MRD) detection, with strong prognostic value demonstrated at the end-of-treatment.^17–20^ Early circulating tumor DNA (ctDNA) kinetics are now being incorporated into response-adapted clinical trials (e.g., NCT04980222).^21,22^ However, these approaches require custom capture panels, paired germline controls, and serial sampling, limiting their adoption.^17,20,23^ Beyond these constraints, MRD status after a single treatment cycle identified a low-risk group. The approximately 30% of patients who achieve MRD clearance after one cycle have an excellent prognosis,^18,24^ making MRD negativity by Phased Variant Enrichment and Detection Sequencing (PhasED-seq) a good candidate for therapy de-escalation in future trials.^18,24^ The challenge lies with the remaining ∼70%, who remain MRD-positive, yet the majority of these patients still achieve favorable long-term outcomes, with a majority of patients remaining progression-free at the 2-year landmark.^24^ Consequently, MRD persistence after one cycle is insufficiently specific to justify therapy escalation, as most MRD-positive patients still do well. Therefore, identifying the high-risk minority within this population requires biomarkers measuring signal beyond tumor burden.

Whole-genome sequencing (WGS) of plasma cfDNA can capture a broader and complementary spectrum of circulating biological signals.^25^ Beyond somatic copy-number aberrations and tumor fraction estimation using tools such as ichorCNA,^26^ low-pass WGS enables analysis of fragment size distributions, fragment-end motif patterns, and regional fragmentation profiles, collectively termed fragmentomics. These features reflect nucleosomal organization, chromatin accessibility, and cell-death pathways in both tumor and non-tumor cells.^25,27–29^ Multimodal integration of these signals has shown additive value in cancer detection frameworks.^27,30^ Fragment-end motif–based approaches such as FrEIA (Fragment-End Integrated Analysis)^30^ and MDS (Motif Diversity Score)^30^ capture cfDNA fragment-end sequence composition and summarize motif diversity which is reflecting enzymatic cleavage of cfDNA. DELFI (DNA EvaLuation of Fragments for early Interception)^31^ evaluates genome-wide cfDNA fragmentation profiles through regional differences in fragment-size distributions. Complementing these fragmentomic approaches, the recently developed LIONHEART (Liquid bIopsy cOrrelatiNg cHromatin accEssibility and cfDNA coverage acRoss cell Types)^32^ software evaluates the cellular sources of cfDNA by correlating the genome-wide cfDNA coverage with known accessible chromatin sites from a large number of non-cancerous and cancerous sources, including cell lines and tissue types. Critically, these approaches are tumor-agnostic, do not require matched germline sequencing, and can be applied to a single sample, properties that make them potentially transformative for early risk stratification in newly diagnosed LBCL, where treatment response reflects both tumor dynamics and systemic biological context.

We hypothesized that integrating genomic and fragmentomic features from a single low-pass cfDNA WGS assay, without tumor tissue or baseline sampling, could enable early and clinically actionable risk stratification in LBCL. In this study, we develop and validate a composite cfDNA score that combines copy-number aberrations and fragmentomic profiles. We benchmark it against established clinical indices and evaluate its potential to identify high-risk patients after just one cycle of therapy to inform future response-adapted treatment strategies.

## Results

### Patients Characteristics and Study Design

We analyzed 288 plasma samples from 190 DLBCL patients and 18 healthy donors (HDs). **Figure 1A** illustrates the study population and sample types. All patients had *de novo* LBCL and were treated with curative-intent immunochemotherapy (see **Methods**). In this cohort (n=190), the median age was 64 years (range, 28-88). Histological subtypes included diffuse large B-cell lymphoma (DLBCL) in 100 patients (52.6%), high-grade B-cell lymphoma (HGBL) with *MYC* plus *BCL2* and/or *BCL6* rearrangements in 89 (46.8%), and primary mediastinal B-cell lymphoma (PMBCL) in 1 (0.5%). Most patients presented with advanced-stage disease (Ann Arbor stage III–IV: 158/190, 83.2%), and International Prognostic Index (IPI) scores were evenly distributed between low-risk (0–2; 94/190, 49.5%) and high-risk groups (3–5; 95/190, 50.0%) (one patient missing IPI) (**Table S1**). Among the 190 patients, 96 had pretreatment samples (T0), 174 had early on-treatment samples collected after one cycle of therapy (T1). For model development and validation, the 174 patients with evaluable T1 samples were divided into a training cohort (Cohort A; n=115; 79 non-progressors, 36 progressors) and an independent validation cohort (Cohort B; n=59; 40 non-progressors, 19 progressors) (**Figure S1**). Progressors (P) were defined as patients who experienced R/R disease within two years of treatment initiation. Non-progressors (NP) achieved complete remission after first-line therapy and remained relapse-free at the two-year landmark. Baseline clinical characteristics including age, stage, and IPI distribution, were well balanced between the training and validation cohorts (**Table S1**).

**Figure 1.**
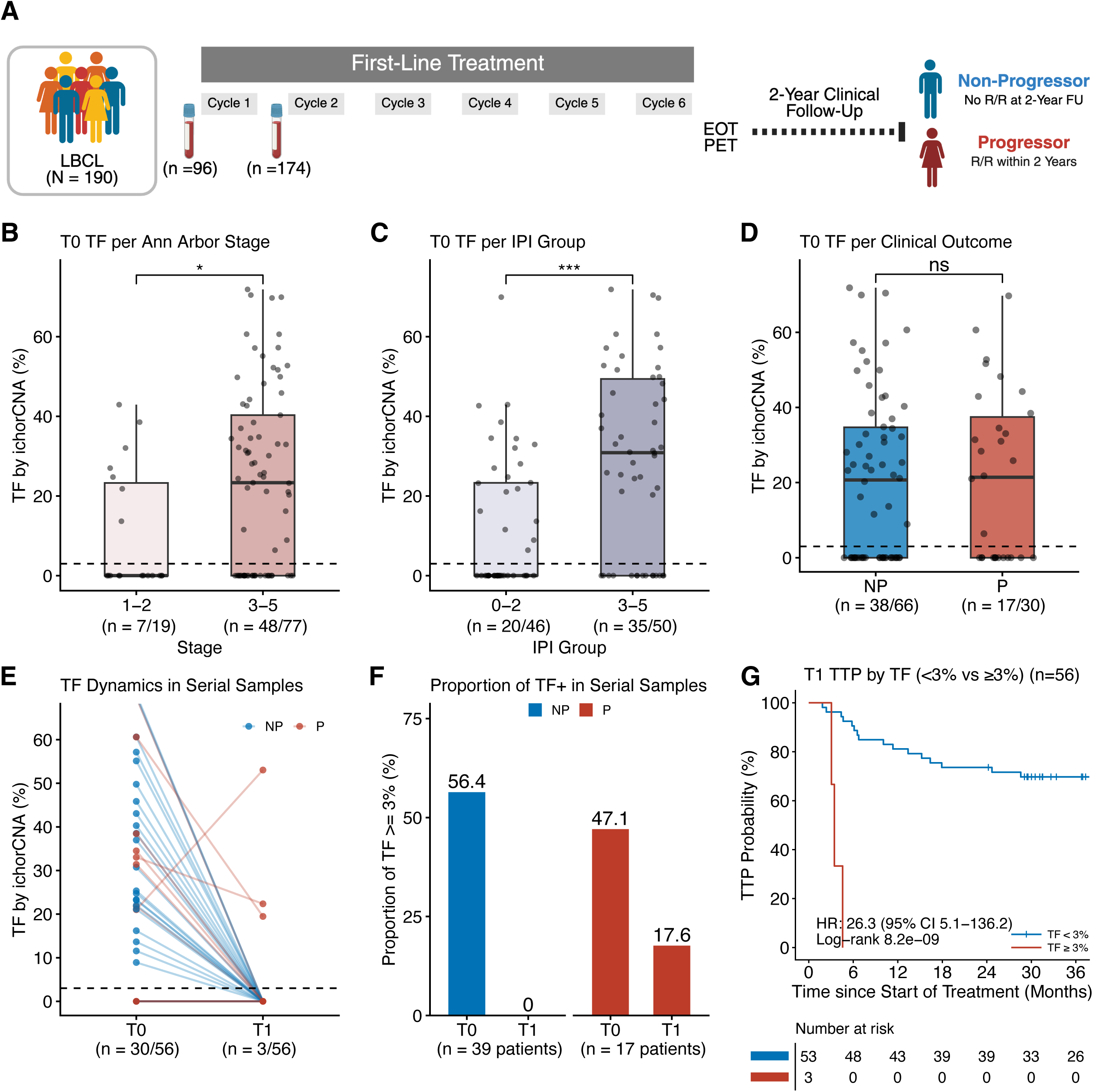
Study design and early on-treatment circulating tumor DNA dynamics. (A) Schematic of the study design. Blood samples were collected in PAXgene circulating cell-free DNA (ccfDNA) tubes from patients with large B-cell lymphoma (LBCL) at baseline (T0) and after one cycle of immunochemotherapy (T1). End-of-treatment PET (EOT PET) served as the standard clinical response assessment following first-line therapy. Non-progressors (NP) were defined as patients who achieved complete remission after first-line therapy and remained relapse-free at the 2-year follow-up (FU), whereas progressors (P) were defined as patients who developed refractory or relapsed (R/R) disease within this period. (B) Baseline tumor fraction (TF), estimated by ichorCNA, stratified by Ann Arbor stage (I-II vs. III-IV). (C) Baseline TF stratified by International Prognostic Index (IPI) (0-2 vs. 3-5). (D) Baseline TF stratified by clinical outcome (NP vs. P). (E) TF dynamics from T0 to T1 in 56 patients in Cohort A with paired samples. The dashed line marks the 3% limit of detection at 95% specificity. (F) Proportion of patients with detectable TF (≥3%) at T0 and T1 in NP and P patients. (G) Kaplan–Meier analysis of time to progression (TTP) stratified by T1 TF status (undetectable, <3% vs. detectable, ≥3%). Hazard ratio (HR) with 95% confidence interval (CI) and log-rank P-value are indicated. Box plots show median and interquartile range (IQR); whiskers extend to 1.5 x IQR. *p < 0.05; **p < 0.01; ***p < 0.001; ns, not significant.

### Early Tumor Burden Divergence between Progressors and Non-Progressors

Tumor fraction (TF) was computed using ichorCNA in all baseline (T0) DLBCL patients (n=96) based on a panel of 18 healthy controls. Following downsampling to 1x coverage, a 3% limit of detection was used.^26^ TF was detectable in 55 of 96 patients (57.2%), with a median TF of 21.1%. While higher stage (**Figure 1B**) and IPI (**Figure 1C**) were associated with higher TF, baseline TF did not differ between NP and P (median TF NP=20.7% vs P=21.4%, *P*=0.9) (**Figure 1D**). We additionally characterized cfDNA using multiple fragmentomic approaches applied to the same sequencing data: fragment size distribution,^33^ LIONHEART,^32^ and DELFI-FTK (DELFI-FinaleToolKit),^34^ which is an executable implementation of the original DELFI approach. At baseline, LBCL patients and HDs differed in fragment size distribution (**Figure S2A**), displayed significant differences in representative LIONHEART features (**Figure S2B**), and showed distinct regional cfDNA fragmentation profiles by DELFI-FTK (**Figure S2C** and **Figure S2D**). Fragment size profiles (**Figure S3A**), LIONHEART features (**Figure S3B**), and DELFI-FTK did not stratify patients by clinical outcome (**Figure S3C** and **Figure S3D**), suggesting that baseline cfDNA whether genomic or fragmentomic, is not sufficient for prognosis.

Then we examined the TF dynamics from baseline (T0) to after one cycle of treatment (T1) in 56 patients in Cohort A with paired samples. After one cycle of treatment, all NP in this subset were TF negative (TF<3%) while three P remained TF positive (TF≥3%) (**Figure 1E**). **Figure 1F** shows the proportion of TF-positive samples at T0 and T1 per clinical outcome. Detection of TF after one cycle of treatment was associated with an inferior prognosis (**Figure 1G**). This motivated us to further explore on-treatment genomic and fragmentomic features.

### On-Treatment Genomic and Fragmentomic Features are Associated with Clinical Outcome

Next, we evaluated genomic and fragmentomic features at T1 in Cohort A (n=115: 79 NP, 36 P). TF calculated with ichorCNA was above detection threshold in 1/79 (1.3%) NP versus 8/36 (22.2%) P (*P*=3.5 10^-4^) (**Figure 2A**). In contrast to baseline, fragmentomic features at T1 diverged significantly between groups. P showed higher proportions of cfDNA fragments in the size range 100-120 bp and 120-140 bp, whereas NP had higher proportions of 180-200 bp fragments (**Figure 2B**). Comparison of trinucleotide fragment-end motifs between P and NP showed that of the motifs enriched in NP, three (CCT, CAT, and CCA) were consistent with trinucleotides previously reported to be depleted in cancer. Conversely, eight of the 12 motifs enriched in P (ACG, GTG, TCG, GAG, AAG, GCG, TTG, and TTC) overlapped with trinucleotides previously reported to be enriched in cancer (**Figure 2C**).^30^ MDS was higher in P than in NP (*P*=2.4 10^-3^) (**Figure 2D**), whereas the FrEIA score did not differ significantly between NP and P (NP vs P; 1.24 vs 1.34, *P*=0.2). Seven LIONHEART features selected by elastic-net feature selection (see **Methods**) significantly differed between P and NP (**Figure 2E, Table S2**). Per-sample mean absolute centered DELFI-FTK was higher in P than NP (Wilcoxon rank-sum test, W=983, *P*=0.014). (**Figure 2F**). Elastic-net selected 77 DELFI-FTK features (see **Methods**, **Table S3**). Features mapping to regions of recurrent copy-number loss in DLBCL, including loci at 6q,^35^ 9p,^35^ 17p,^35^ and 8p^36^, formed a broadly coherent signal, with most contributing in the expected direction. Features corresponding to established oncogenic amplicons at 3q,^35^ 18q21,^35^ 2p16,^35^ and 8q^35^ also ranked among the most important. The general correspondence between the highest-ranked DELFI-FTK features and previously reported cytogenetic markers in LBCL suggests that the fragmentomic signal captures, at least in part, biologically relevant aspects of disease. Together, these results indicate that both genomic and fragmentomic profiles stratify patients by outcome after one cycle of immunochemotherapy, supporting their potential utility for early response assessment and predictive modeling.

**Figure 2.**
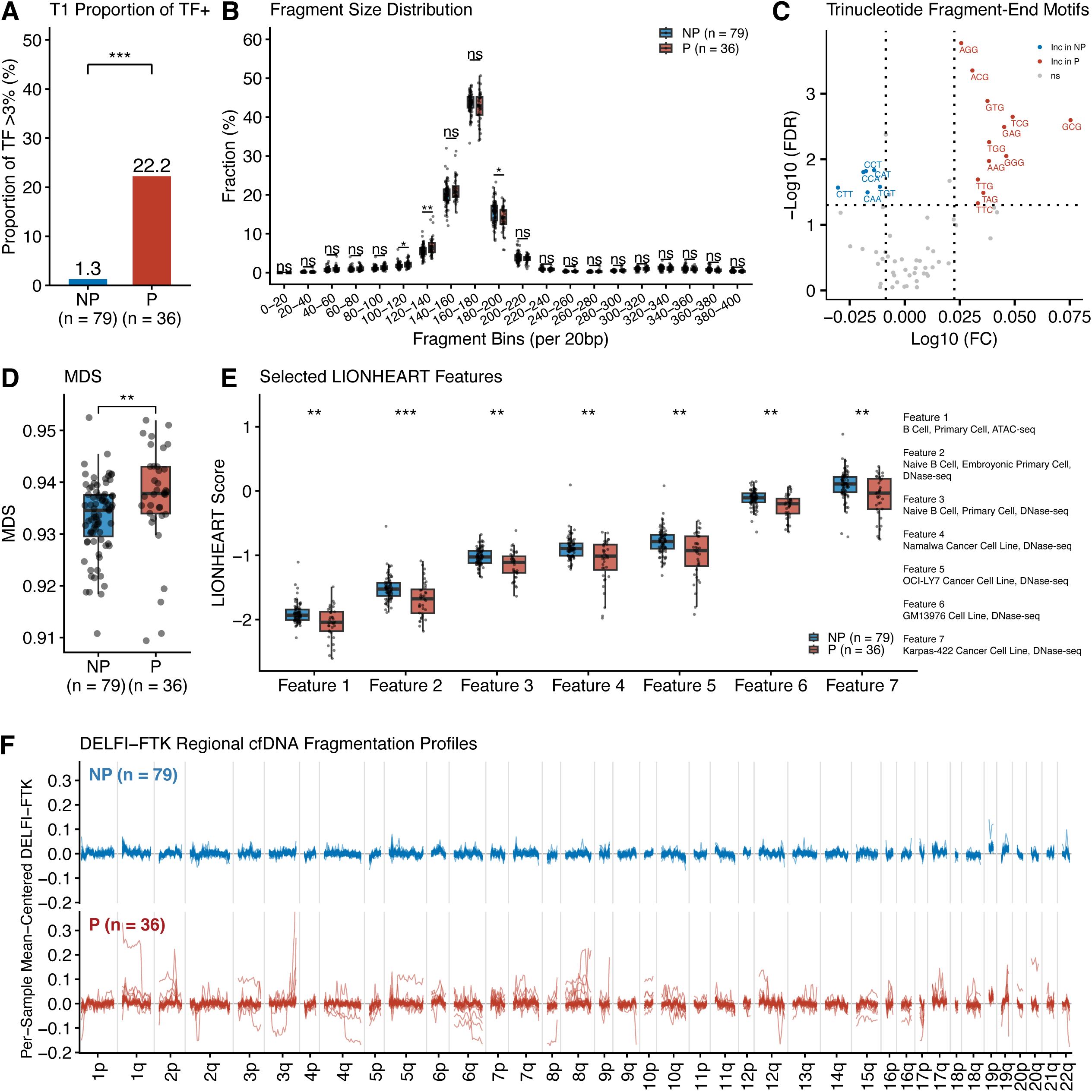
On-treatment genomic and fragmentomic features distinguish non-progressors from progressors in Cohort A. (A) Proportion of TF-positive T1 samples per clinical outcome. (B) Fragment size distributions at T1 (after one cycle of immunochemotherapy) in non-progressors (NP) and progressors (P), shown in 20 bp bins. (C) Trinucleotide fragment-end motifs enriched in NP and P patients at T1. (D) Motif Diversity Score (MDS) at T1 compared between NP and P patients. (E) Selected LIONHEART features at T1 in NP and P patients. (F) DELFI-FTK regional cfDNA fragmentation profiles across chromosome arms by clinical outcome at T1. Box plots show median and interquartile range (IQR); whiskers extend to 1.5 x IQR. *p < 0.05; **p < 0.01; ***p < 0.001; ns, not significant.

### ACT Score: A Multimodal cfDNA Model for Outcome Prediction

To evaluate whether a combination of cfDNA features could improve prediction of 2-year time-to-progression (TTP), we examined the intercorrelations among four genomic and fragmentomic features not selected by Elastic Net, namely TF, fragment size, FrEIA, and MDS (**Figure S4**). At T1, all cross-modality correlations with |ρ|≥0.3 were statistically significant, but none exceeded ∼0.4, remaining below the conventional redundancy threshold of 0.7–0.8. TF showed moderate correlations with MDS (|ρ|=0.35), FrEIA (|ρ|=0.31), and the fragment bins (|ρ|=0.30–0.40), consistent with tumor burden contributing to shorter and more aberrant cfDNA fragments; yet 80–90% of the variance in each fragmentomic feature remained independent of TF. MDS and FrEIA were only weakly correlated with each other (ρ=−0.21), suggesting that they capture distinct aspects of cfDNA biology. Together, these findings indicate that TF, fragment-size summaries, MDS, and FrEIA act as complementary predictors (**Figure S4**).

We developed the ACT score (Aberrations, Composition of fragments, and Terminal motif analyses) in the training cohort by using logistic regression to integrate selected genomic and fragmentomic features derived from a single cfDNA sequencing run and subsequently evaluated its performance in the validation cohort (**Figure 3A**). The ACT score incorporated genomic and fragmentomic features, including TF, fragment size distributions across three 20 bp bins, MDS, FrEIA, and selected LIONHEART and DELFI-FTK features (see **Methods**). Hyperparameter ranges evaluated during model optimization are shown in **Table S4**. An optimized classification threshold of 0.54, derived in Cohort A as described in the Methods, was applied to Cohort B to classify patients as ACT-positive or ACT-negative for validation. **Table S5** lists the features and corresponding coefficients from the final logistic regression model. Positive coefficients increase the predicted probability of progression within two years, whereas negative coefficients decrease this probability.

**Figure 3.**
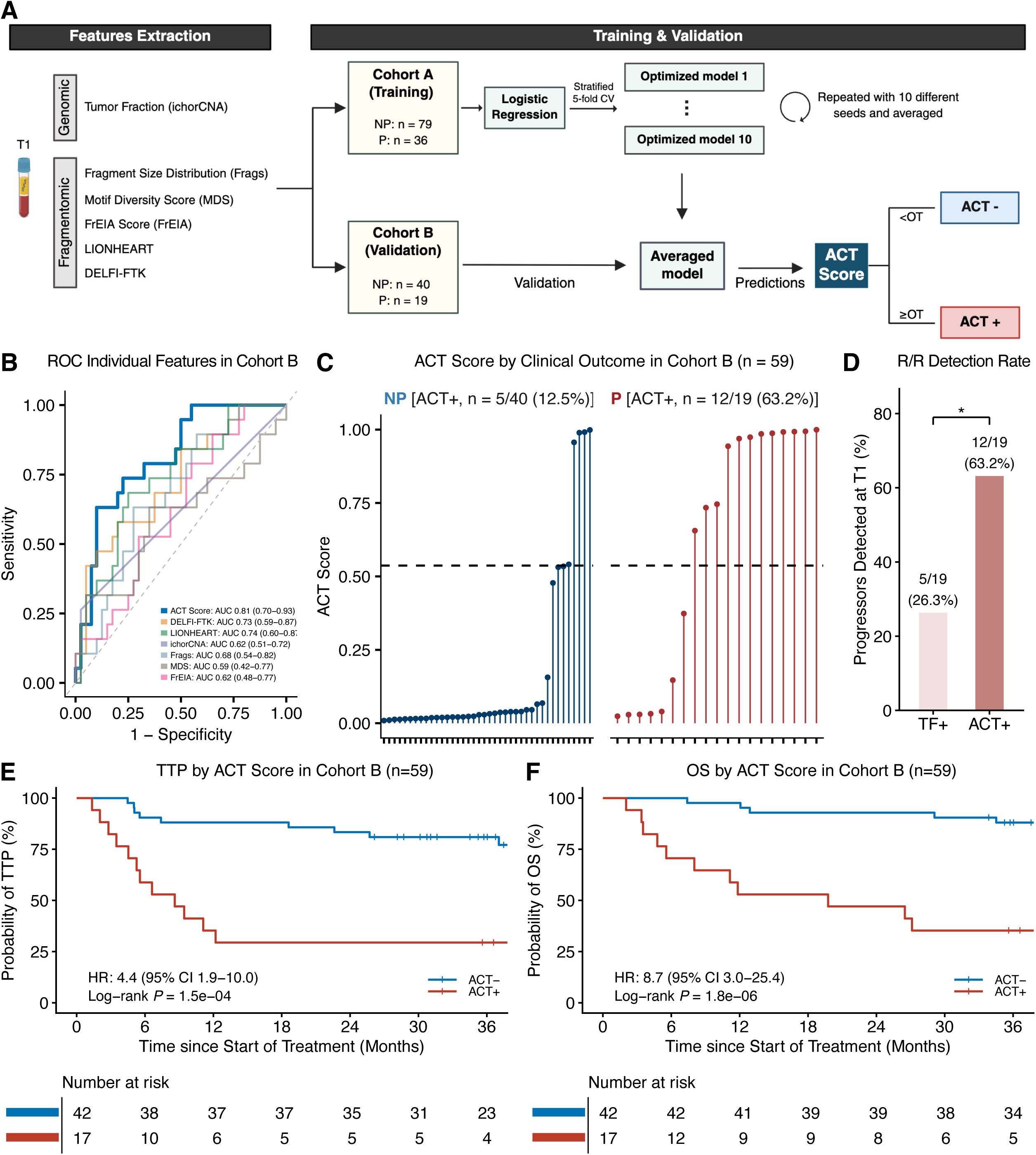
Development and independent validation of the ACT score for early prediction of treatment outcome. (A) Machine-learning workflow for ACT (Aberrations, Composition of fragments, Terminal motif analyses) score development and validation. A logistic regression model was trained in Cohort A (training cohort) to predict 2-year time-to-progression (TTP) using six cfDNA feature sets. Hyperparameters and the classification threshold (optimal threshold, OT) were optimized by stratified 5-fold cross-validation (CV). To ensure robustness, model training was repeated with 10 random seeds, yielding 10 optimized models, each generating an ACT score. These scores were then averaged to produce the final predictions. In the independent validation cohort (Cohort B), the averaged ACT scores and classification threshold were used to classify samples as ACT+ or ACT-. (B) Receiver operating characteristic (ROC) curves of the ACT score and individual genomic and fragmentomic features for predicting 2-year TTP. The area under the ROC curve (AUC) for the ACT score and each individual feature is shown with 95% confidence intervals (CI). (C) Lollipop plots of individual ACT scores in Cohort B. Each lollipop represents an individual patient, colored by clinical outcome (blue, NP; red, P). Points indicate the mean score across ten iterations. An optimized threshold of 0.54 stratifies patients into predicted progressors (ACT+, ACT ≥ 0.54) and predicted non-progressors (ACT−, ACT < 0.54). (D) Detection rate (sensitivity) for progressors in Cohort B, comparing classification based on T1 tumor fraction (TF) alone with classification based on the ACT score. TF+ was defined as TF ≥ 3%. (E) Kaplan–Meier estimates of TTP in Cohort B, stratified by ACT score. (F) Kaplan–Meier estimates of overall survival (OS) in Cohort B, stratified by ACT score. Hazard ratio (HR) with 95% CI and log-rank *P*-value are indicated. *p < 0.05; **p < 0.01; ***p < 0.001; ns, not significant.

In Cohort B, the ACT score achieved an area under the ROC curve (AUC) of 0.81 (95% CI: 0.70-0.93) for predicting 2-year TTP and outperformed the individual features included in the model development (**Figure 3B**). **Figure 3C** shows Individual patient predictions per clinical outcome. ACT score derived using logistic regression outperformed five other benchmarked machine learning models with a sensitivity of 63%, specificity of 88%, positive predictive value (PPV) of 71%, and negative predictive value (NPV) of 83% (**Table S6**). **Table S7** provides a complete overview of clinical characteristics, individual genomic and fragmentomic features, and predicted versus actual clinical outcome of all patients. Descriptions of the variables included in Table S7 can be found in **Table S8**. At T1, the ACT score identified 12 of 19 progressors (63%) in the validation cohort, compared to 5 of 19 (26%) identified by TF alone (**Figure 3D**), an absolute improvement of 37%, demonstrating that fragmentomic features capture prognostically relevant signals that are complementary to tumor burden. The ACT score yielded a higher AUC than tumor fraction alone for the 24-month landmark (0.81 vs 0.62; DeLong P=5 × 10^-4^). Patients who were ACT-positive had significantly inferior survival outcomes: 2-year TTP was 29% compared to 83% in ACT-negative patients with an hazard ratio (HR) of 4.4 (95% confidence interval [CI]: 1.9-10.0, Log-rank *P*=1.5 × 10^-4^) (**Figure 3E**), and 2-year OS was 47% versus 93%, HR 8.7; 95% CI: 3.0–25.4; Log-rank *P*=1.8 × 10^-6^) (**Figure 3F**). The prognostic value of the ACT score for TTP and OS was consistent across histological subtypes, with significant separation observed in both DLBCL and HGBL subgroups (**Table S9**). Together, these results demonstrate that combining cfDNA genomic and fragmentomic features from a single plasma sample after one cycle of immunochemotherapy enables early prediction of long-term clinical outcomes.

### ACT Score Complements IPI and iPET-CT for Outcome Prediction

To assess the prognostic value of the ACT score in the context of existing clinical risk stratification tools, we compared its performance against the IPI and interim PET (iPET) performed after three cycles of treatments. Performance of IPI and iPET for predicting TTP and OS in all patients where data are available are shown in **Figure S5**. In the validation cohort (Cohort B), IPI scores were available for 58 of 59 patients. IPI was not predictive of TTP (HR 2.2 95% CI 0.9–5.3, Log-rank *P*=0.068) (**Figure 4A**) but for OS (HR 3.1 95% CI 1.0–9.5, Log-rank *P*=0.041) (**Figure 4B**). When combined with IPI, patients with low IPI and negative ACT score had the lowest risk of relapse (2-year TTP 89%), while all IPI high-risk and ACT-positive patients experienced R/R disease within two years (Log-Rank *P*=1.1 × 10^-9^) (**Figure 4C**). The combination of IPI and ACT was also prognostic for OS (Log-Rank *P*=9.4 × 10^-15^) (**Figure 4D**). Multivariate analysis showed that ACT score remained independently prognostic for TTP (**Figure 4E**) (HR 7.0 95% CI 2.8-17.5, *P*=3.6 × 10^-5^) and OS (HR 23.5 95% CI 6.5-85.5, *P*=1.6 × 10^-6^) (**Figure 4F**) when adjusted for IPI. In the subset of Cohort B patients with iPET available after cycle three (n=32), iPET alone and in combination with the ACT score was prognostic for both TTP and OS (**Figure S6A-D**). ACT score was independently prognostic for TTP and OS when adjusted for iPET (**Figure S6E**). In this subset, patients with low IPI, ACT negativity, and negative iPET had 100% 2-year TTP. These findings suggest that baseline IPI and ACT score may help identify high-risk patients, enabling therapy escalation from cycle three. Conversely, no patients with low-risk IPI, ACT negativity, and negative iPET experienced progression by the 2-year landmark.

**Figure 4.**
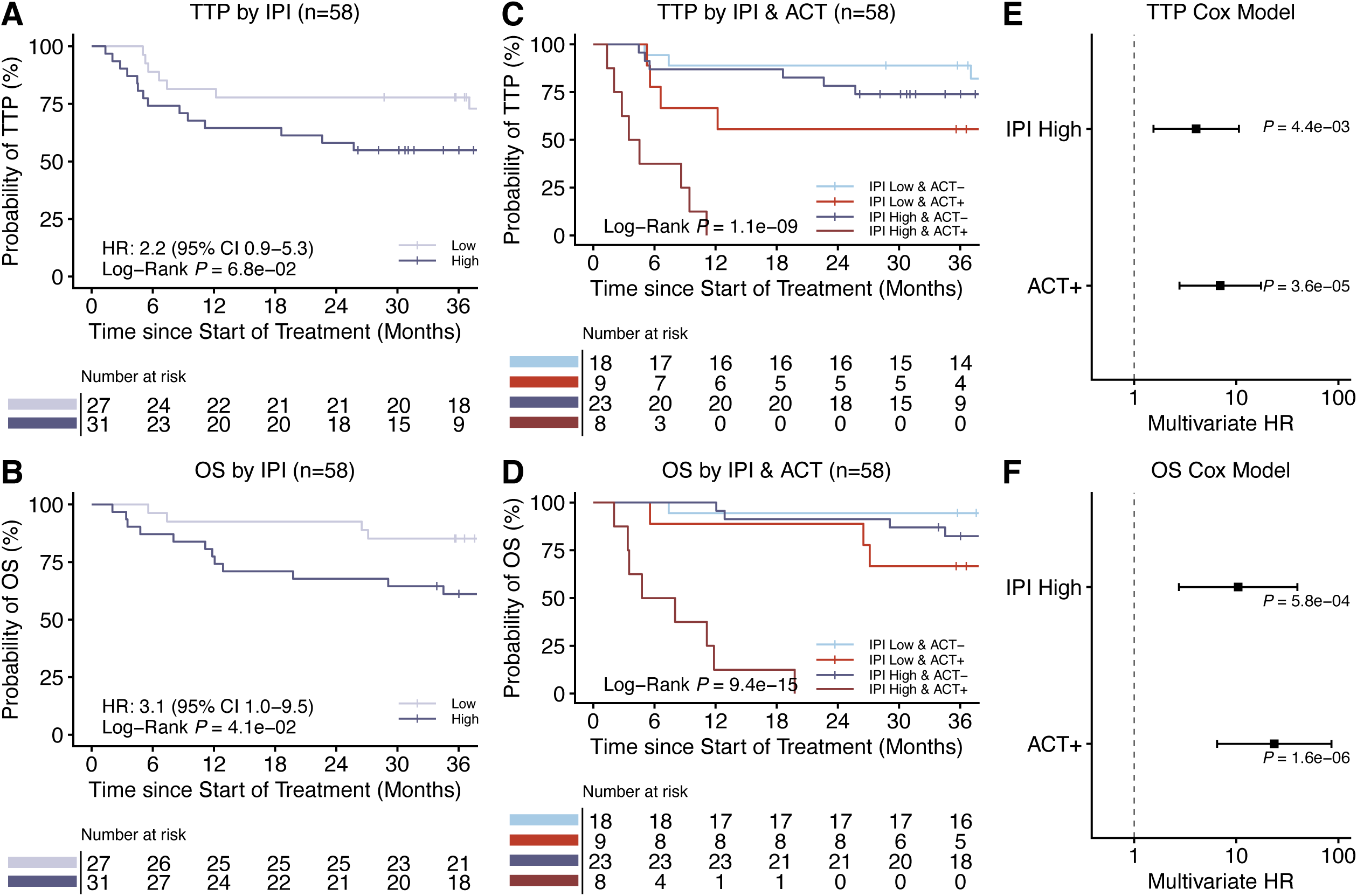
ACT score provides independent prognostic value beyond IPI. (A) Kaplan–Meier estimates of time-to-progression (TTP) stratified by International Prognostic Index (IPI; 0–2 vs. 3–5) in patients with both IPI and ACT score available in Cohort B (n=58). (B) Kaplan–Meier estimates of overall survival (OS) stratified by International Prognostic Index (IPI; 0–2 vs. 3–5) in patients with both IPI and ACT score available in Cohort B (n=58). (C) Kaplan–Meier estimates of TTP stratified by combined ACT score and IPI in patients with both metrics available in Cohort B (n=58). (D) Kaplan–Meier estimates of OS stratified by combined ACT score and IPI in patients with both metrics available in Cohort B (n=58). (E) Forest plot of multivariable Cox proportional hazards analysis for TTP in Cohort B, including IPI (3-5) and ACT score (ACT+) as covariates. (F) Forest plot of multivariable Cox proportional hazards analysis for OS in Cohort B, including IPI (3-5) and ACT score (ACT+) as covariates. Hazard ratio (HR) with 95% CI and log-rank *P*-value are indicated. *p < 0.05; **p < 0.01; ***p < 0.001; ns, not significant.

## Discussion

We present a minimally invasive, open-access, and low coverage WGS-based strategy for early risk stratification in patients with newly diagnosed LBCL. Using plasma cfDNA collected after one cycle of immunochemotherapy, without the need for tumor tissue, germline controls, or baseline samples, we developed the ACT score by integrating six complementary genomic and fragmentomic features. ACT score identifies patients at high risk of treatment failure early during therapy. In the independent validation cohort, ACT-positive patients had substantially worse outcomes than ACT-negative patients, with a two-year TTP of 29% versus 83% and a two-year OS of 47% versus 93%, respectively. The prognostic effect of ACT exceeded that of the IPI and remained independent of IPI in multivariable analysis. When combined with IPI, the ACT score identified a high-risk subgroup who may benefit from therapy escalation, while ACT-negative patients with high IPI retained a favorable prognosis.

The ACT score is different from targeted ctDNA assays designed for MRD detection. Tumor-informed methods such as PhasED-Seq achieve high analytical sensitivity through mutation tracking across serial samples, but require matched germline DNA, custom sequencing panels, and repeated sampling, barriers that limit accessibility beyond the cost in many clinical settings.^37,17,38,20^ By contrast, the ACT score is tumor-naive, applied at a single timepoint, and built on open-source tools deployable on standard WGS platforms and data. This distinction also clarifies the clinical questions each approach answers. MRD and tumor-guided assays are best suited for remission assessment and therapy de-escalation in patients achieving deep molecular responses in hematological malignancies.^39,40^ Recent data indicate that ∼70% of patients who remain MRD-positive with phased variant technologies after one cycle are still relapse-free at three years,^18,24^ meaning MRD persistence at this early timepoint is insufficiently specific to justify systematic therapy escalation. By contrast, the ACT score is designed to support treatment-escalation decisions by identifying the subset of patients, including approximately two-thirds of eventual progressors, for whom treatment intensification may be most warranted. This ability stems from the integration of both tumor burden and broader biological context in the ACT score currently missed by mutation-based MRD assays like PhasED-Seq. The combination of IPI and ACT identified two distinct patient groups: low-risk (IPI 0–2 and ACT−) and high-risk (IPI 3–5 and ACT+), reflecting the potential of using these biomarkers to guide treatment decisions from cycle 3 onward (Figure 5). A sub-analysis of iPET performed after three cycles showed that iPET-negative patients had excellent outcomes. Since iPET-guided therapy has become standard of care in the Netherlands, this approach could be leveraged to determine whether low-risk patients (IPI 0–2 and ACT−) who are also iPET-negative after cycle 3 may safely discontinue R-CHOP after four cycles, followed by two additional cycles of rituximab.

**Figure 5.**
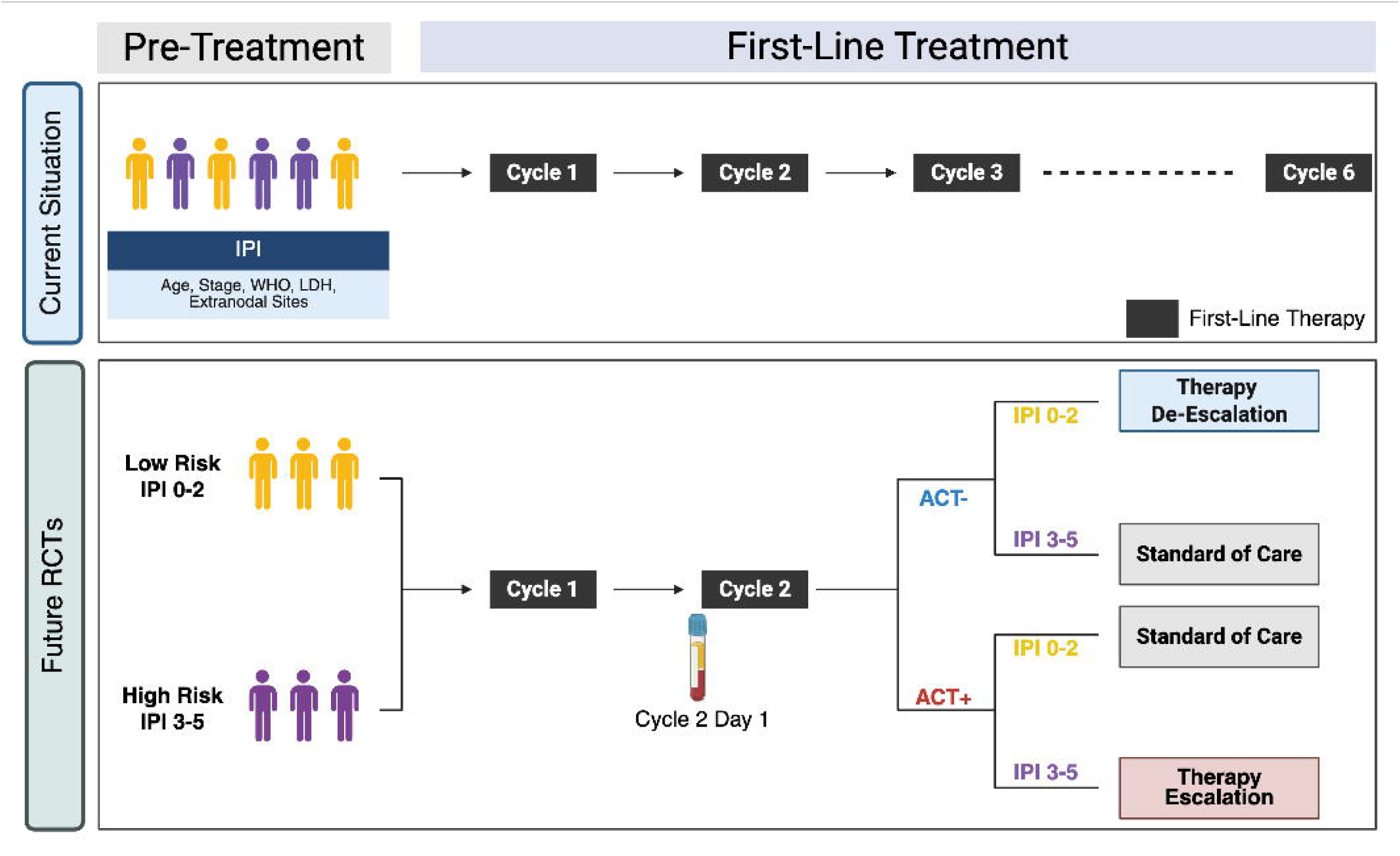
ACT score in combination with IPI has the potential to support future interventional trials. Pre-treatment risk stratification using the International Prognostic Index (IPI) is suboptimal for identifying patients who will or will not benefit from standard first-line treatment. In the proposed liquid biopsy–based strategy, all patients initiate standard R-CHOP, and peripheral blood is collected on cycle 2 day 1 (i.e., after a single cycle of therapy) for multi-modal cell-free DNA analysis. ACT score enables response-adapted treatment modification beginning at cycle 3. ACT score can be used, in combination with IPI, to identify low-risk (IPI 0-2 & ACT-) and high-risk (IPI 3-5 & ACT+) patients, providing opportunities for future therapy de-escalation and escalation trials. Figure icons represent individual patients; colors denote distinct risk groups, illustrating that ACT score, rather than baseline IPI alone, drives treatment adaptation. RCT: Randomized Clinical Trials.

Recent reports have highlighted the potential of multimodal combinations across a range of liquid biopsy applications,^27,30,41,42^ underscoring that distinct cfDNA signals capture biologically non-redundant information: some reflect tumor-derived biology, whereas others capture non-tumor compartments and host responses. Notably, after just one cycle of treatment, non-progressors more closely resembled HDs, whereas progressors more closely resembled baseline LBCL samples in terms of fragment size distribution.^30^ In our study, analysis of trinucleotide fragment-end motifs after one cycle yielded similar findings,^30^ further supporting the notion that orthogonal fragmentomic features converge on the same early-response signal. LIONHEART features with lower scores in progressors, indicating greater contribution from the corresponding cell or tissue types, included both malignant B-cell and non-malignant B-cell signals, consistent with LBCL biology and the higher TF observed in progressors. DELFI-FTK features associated with progression included regions on chromosome arms 9p and 17p, which have been previously linked to poor clinical outcomes.^43,44^ Tumor burden was captured directly by ichorCNA,^26^ and indirectly by both FrEIA^30^ and the OCI-Ly7/B-cell LIONHEART^32^ features, collectively suggesting that higher levels of tumor-derived cfDNA are associated with worse outcome. The alignment of the highest-ranked DELFI-FTK features with genomic regions harboring well-characterized recurrent deletions and oncogenic amplicons in LBCL lends biological plausibility to the fragmentomic signal and suggests that sWGS of cfDNA may, at least partly, reflect the underlying tumor cytogenetic landscape. Achieving this without targeted sequencing or prior knowledge of tumor-specific alterations highlights the potential breadth of genome-wide fragmentomic approaches for capturing clinically relevant disease biology from plasma. Cell-of-origin and microenvironmental signals also emerged from the LIONHEART features, which are based on cfDNA coverage patterns linked to chromatin accessibility across tumor and non-tumor reference cell types and may reflect aspects of host immune composition. However, the direction of individual cell-type associations should be interpreted cautiously because of feature collinearity. Together, the convergence of tumor-intrinsic and non-tumor signals across these blocks illustrates why multimodal cfDNA profiling may add value beyond any single assay.

Early identification of high-risk patients likely to experience R/R disease is essential for risk-adapted treatment strategies in LBCL. Conventional prognostic tools such as the IPI and COO classification are limited in their ability to identify high-risk patients.^14^ Prior efforts on quantification of pretreatment ctDNA concentration in human genome equivalent per mL (hGE/mL) has shown that high pretreatment ctDNA concentration is associated with an inferior outcome,^21,45–47^ however these studies use different panels and thresholds, and label up to nearly 50% of patients as high-risk. Despite dichotomous baseline ctDNA concentration achieving statistical significance for predicting clinical outcome, this method would flag nearly half of all patients for therapy escalation, raising safety concerns about overtreatment. As a result, on-treatment response assessments are emerging as more reliable predictors for risk-adapted therapeutic strategies. Molecular response after one cycle has shown strong prognostic value for event-free survival (HR=11.5, p=0.0047) and already guiding therapy escalation in an ongoing clinical trial using the AVENIO Oncology Assay for Non-Hodgkin’s Lymphoma (NCT04980222).^21,22^ Our findings are consistent with this paradigm: the ACT score was associated with long-term outcome as early as after one treatment cycle. These results suggest that the ACT score could be integrated into future risk-adapted treatment strategies, pending further validation. Notably, this approach parallels the model established by non-invasive prenatal test (NIPT), which also employs WGS and has been successfully implemented at scale in several European and Asian countries.^48,49^ The existing infrastructure supporting NIPT, including technical pipelines and regulatory approvals, provides a strong precedent for deploying similar WGS-based cfDNA assays in oncology.

In short, risk-adapted treatment strategies for patients with LBCL depends on biomarkers that address different clinical needs.^14^ Therapy de-escalation would likely require tumor-specific MRD techniques as recent data showed that up to 30% of patients who achieved MRD clearance after one cycle of treatment had no relapse for three years from treatment initiation.^18,24^ However, the remaining ∼70% still had a relatively favorable prognosis, with more than 60% remaining free from progression at 2 years, making MRD clearance after one cycle unsuitable for identifying patients for therapy escalation. By contrast, ACT score identifies nearly two-thirds of eventual progressors and can be used in combination with IPI. **Figure 5** illustrates one possible framework for risk-adapted treatment in LBCL.

### Limitations of the study

Nonetheless, several limitations warrant consideration when interpreting the results. Firstly, our cohorts included patients treated in two prospective trials with distinct regimens: DLBCL-NOS patients received R-CHOP, while HGBL-DH/TH patients were treated with one cycle of R-CHOP followed by DA-EPOCH-R and consolidation with nivolumab in patients achieving complete metabolic response. Despite the difference in regimen from cycle two, the superiority of DA-EPOCH-R over R-CHOP has never been prospectively demonstrated for HGBL-DH/TH patients. While the distribution of patients receiving nivolumab consolidation was similar across the training and validation cohorts, and while the clinical benefit of checkpoint inhibitor consolidation remains debated,^50^ the effects of these regimens on outcome may not be equivalent. Fragmentomic features may be less tumor-specific than genomic alterations, as they can be influenced by broader biological processes, including cell death and immune perturbations.^51^ Future studies should prospectively evaluate the ACT score in treatment-homogeneous cohorts to disentangle regimen-specific effects. Secondly, the shallow depth of WGS used in this study does not enable reliable detection of point mutations or structural variants. However, as high-depth WGS becomes more affordable with the emergence of new sequencing technologies, and as analytical pipelines continue to mature, genome-wide cfDNA profiling may serve both as an early risk-stratification tool and as a bridge to more targeted, sensitive downstream molecular assays.^27,41,52–54^ Higher-depth sequencing could enable the capture of more diverse and granular fragmentomic signals, further enhancing the potential of the ACT score.^29^ In parallel, emerging chemistries may expand the liquid-biopsy signal space to include underexplored features such as cfDNA fragment jaggedness and topology. This potential could be further enhanced by emerging multimodal chemistries that enable the simultaneous capture of genomic and epigenetic alterations within a single sequencing experiment.^55^ Third, whether fragmentomic and genomic features exhibit similar or divergent trajectories between cycles was not assessed, as this is beyond the scope of our primary objective of early risk stratification; such analyses represent an interesting avenue for future investigation. Finally, the findings of this study warrant validation in a larger cohort before supporting future interventional trials.

In conclusion, the ACT score represents an open-source and easily standardizable minimally-invasive method for early outcome prediction in patients with aggressive B-cell lymphomas. By integrating genomic and fragmentomic cfDNA features from a single on-treatment plasma sample, this approach offers a clinically feasible alternative to mutation-based methods for risk stratification and may enable timely risk-adapted treatment decisions. Future randomized trials are needed to evaluate the utility of ACT-guided therapeutic strategies, such as early escalation to bispecific antibodies or transition to CAR T-cell therapy in patients with poor early molecular response.

## Data Availability

Sequencing data generated in this study have been deposited in the European Genome-Phenome Archive (EGA) under accession numbers EGAD50000000602.

## Resource availability

### Lead contact

Requests for further information should be directed to the lead contact, Florent Mouliere.

### Data and code availability

- Data: Sequencing data generated in this study have been deposited in the European Genome-Phenome Archive (EGA) under accession numbers EGAD50000000602. Anonymized clinical data has been reported in Supplemental Information.
- Code: Machine learning models used for feature selection and constructing the ACT-score have been deposited on GitHub.

## Acknowledgments

The authors thank all participants of the HOVON trials for their contributions to this study. We are grateful to the Clinical Genetics facilities at Amsterdam UMC for their support, and to the Amsterdam UMC Liquid Biopsy Center for their logistical assistance and valuable input. The authors would like to extend our gratitude to Professor Idris Bahce, MD/PhD, who provided healthy donor samples through the Liquid Biopsy Center. The authors thank Dr. Steven Hill and Dr. Chao Han from the Cancer Research UK (CRUK) National Biomarker Centre for supporting machine learning analysis. Y.P. and F.M. were supported by the Amsterdam UMC Liquid Biopsy Center, an initiative made possible through the Cancer Center Amsterdam. This work was carried out in part on the Dutch national e-infrastructure with support from the SURF Cooperative. The authors also acknowledge the Netherlands Comprehensive Cancer Organization (IKNL) and PALGA for their collaboration and data support. M.E.D.C. and S.W. gratefully acknowledge support from the Fentener van Vlissingen Foundation. F.M. is funded by CRUK via core funding to the CRUK National Biomarker Centre and supported by the Manchester Experimental Cancer Medicines Centre, the CRUK Lung Cancer Centre of Excellence, and the NIHR Manchester Biomedical Research Centre.

## Authors contribution

Conceptualization, S.W., P.M., N.M., Y.P., M.E.D.C., and F.M.; Methodology, S.W., P.M., N.M., Y.P., and F.M.; Investigation, S.W., Y.P., N.M., P.M., A.D.B.V, N.A.T., B.S., A.S., and F.M.; Clinical data management, E.W., A.D.; Analysis, S.W., Y.P., N.M., P.M., F.M., and A.S.; Sample acquisition, S.W., A.V.J., E.E.E.D., M.N., M.P., K.H., C.K., R.R., R.F., V.K.J.V., A.B., L.N., R.M., and J.S.P.V., M.E.D.C.; Funding acquisition, D.M.P., M.E.D.C., and F.M.; Writing – original draft, S.W., P.M., Y.P., and F.M.; Writing – review & editing, all authors; Supervision, D.M.P., M.E.D.C., and F.M.

## Declaration of Generative AI and AI-assisted Technologies in the Writing Process

During the preparation of this work the author(s) used ChatGPT 5.5 by OpenAI and Claude Opus 4.7 by Anthropic to improve the readability of the manuscript and the generation of the graphical abstract that was refined by the authors. After using these tools, the authors reviewed and edited the content as needed and take full responsibility for the content of the published article.

## STAR Methods

### Key resources table

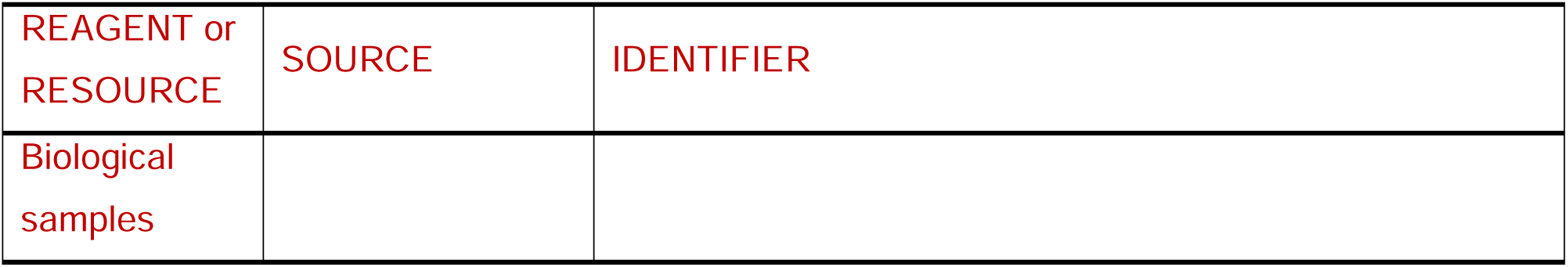
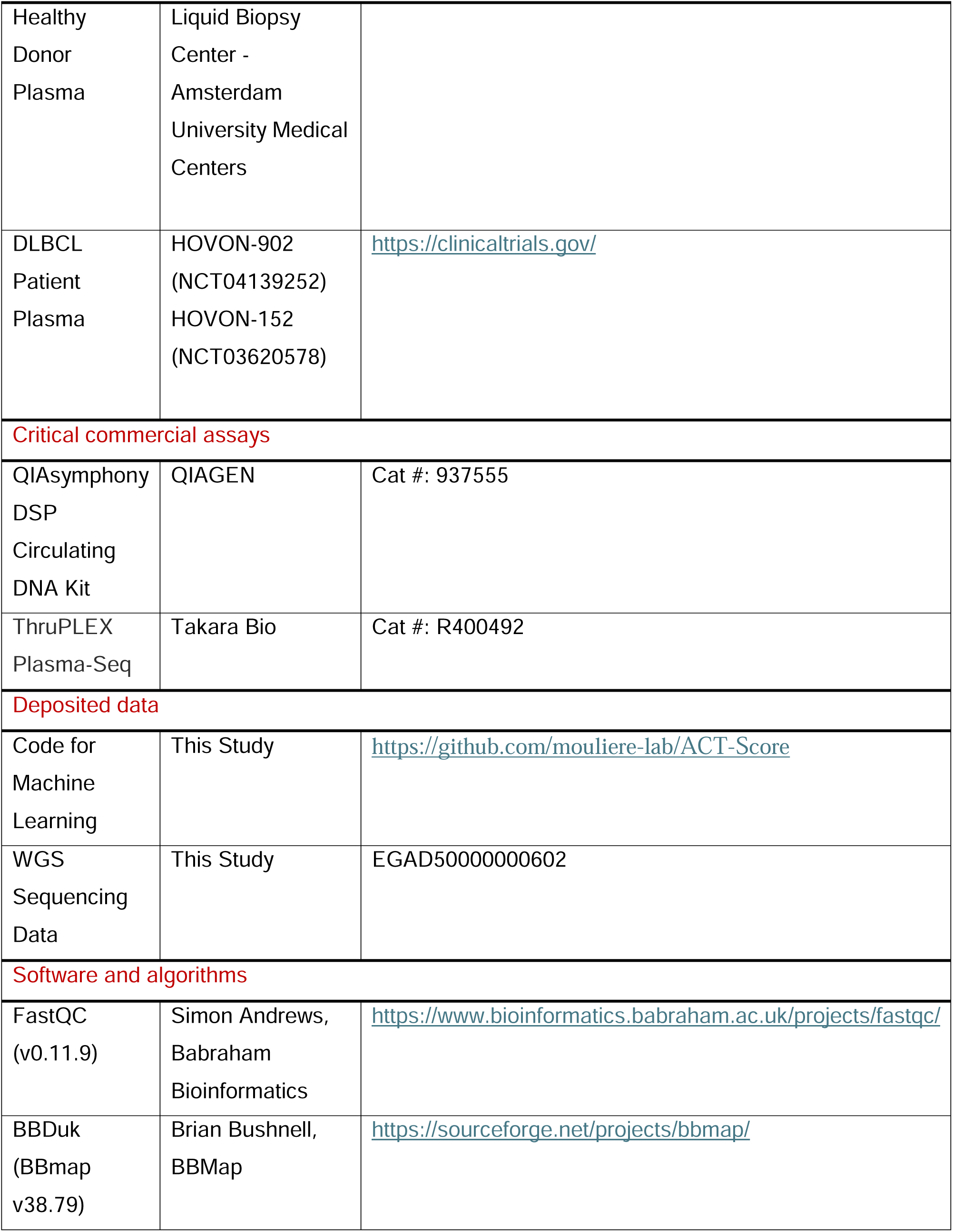
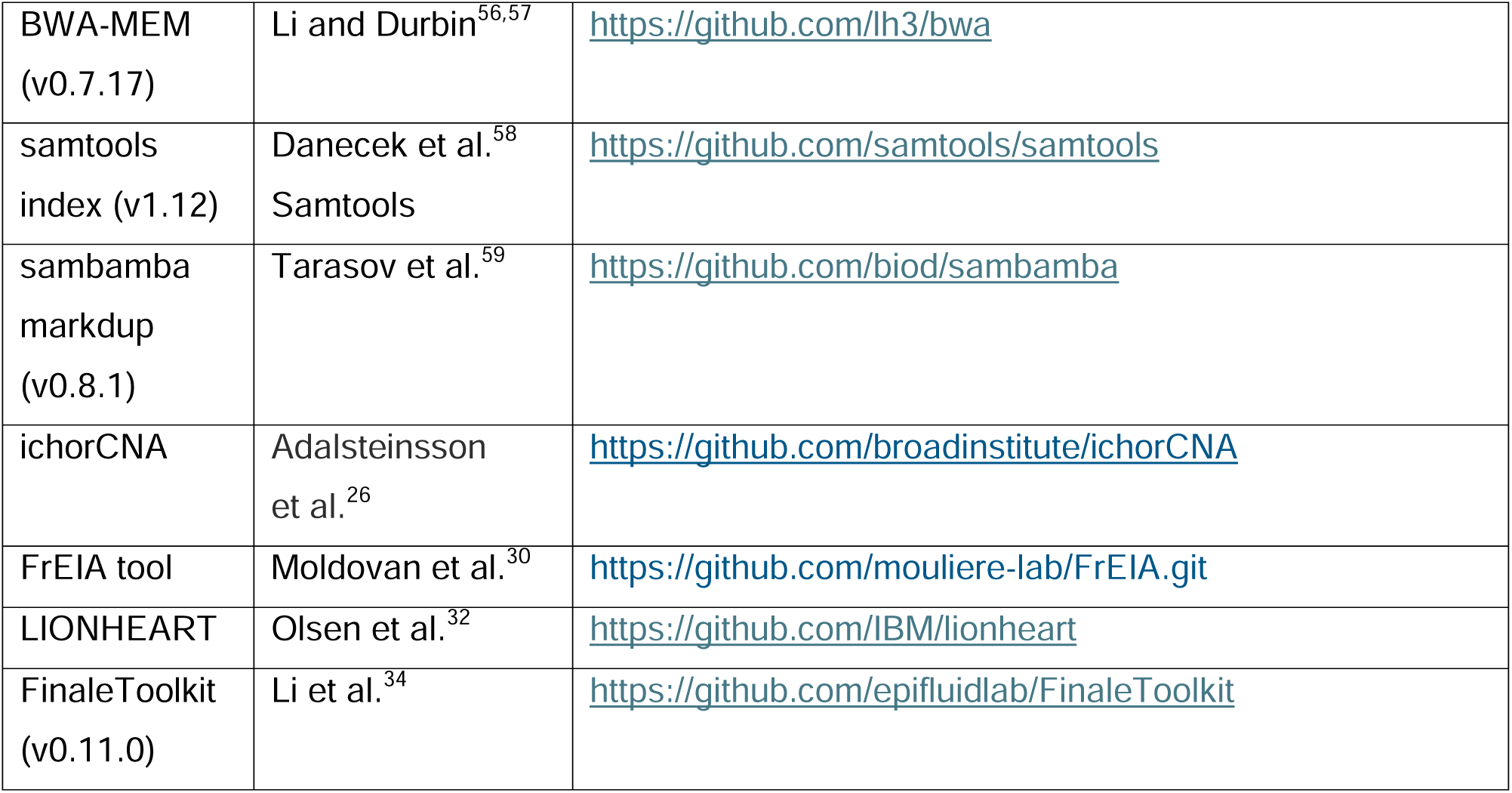

### Experimental model and study participant details

#### Human samples

We analyzed plasma from newly diagnosed DLBCL patients (DLBCL-NOS, HGBL-NOS, and HGBL-DH/TH per 2016 WHO criteria) enrolled in two Dutch-Belgian multicenter studies: HOVON-902 (NCT04139252), an observational biomarker study across 55 centers in patients receiving six cycles of R-CHOP or DA-EPOCH-R; and HOVON-152 (NCT03620578), a single-arm Phase 2 trial for HGBL-DH/TH patients treated with one cycle of R-CHOP followed by five cycles of DA-EPOCH-R, with one-year nivolumab consolidation for those achieving complete metabolic response. Response was assessed by PET-CT after the final immunochemotherapy cycle using Deauville criteria, with interim PET-CT after cycle three. Both studies were approved by the Institutional Review Board (NL70323.029.19 and NL63247.029.17), conducted per the Declaration of Helsinki, and enrolled patients between 2018 and 2022. HD plasma samples were obtained from the Amsterdam UMC Liquid Biopsy Center.

### Method details

#### Study Design

In total, we analyzed 299 samples from 196 newly diagnosed LBCL patients and 18 healthy donors (HDs). Ten samples from six patients were excluded due to COVID-19-related death or second malignancies. One sample was excluded based on quality control (QC) flagging for possible mislabeling. As a result, 288 plasma samples obtained from 190 patients and 18 healthy controls. Samples were collected at baseline and after one cycle of immunochemotherapy. From these samples, we extracted six cfDNA-derived features: (1) Tumor Fraction (TF), (2) fragment size profiles, (3) Motif Diversity Score (MDS), (4) Fragment End Integrated Analysis (FrEIA) score, (5) Liquid bIopsy cOrrelatiNg cHromatin accEssibility and cfDNA coverage acRoss cell Types (LIONHEART), and (6) DNA EvaLuation of Fragments for early Interception (DELFI-FTK). We assessed the prognostic value of each individual feature and subsequently integrated them into a multi-modal classifier. To build and validate the model, samples were divided into a training cohort (Cohort A, n=115) from 27 hospitals and an independent external validation cohort (Cohort B, n=59) from 15 additional centers (**Figure S1**).

#### Sample Processing

Peripheral blood was collected in 10 mL PAXgene Blood circulating cell-free DNA (ccfDNA) Tubes (QIAGEN, Germany) and processed centrally within 48 hours. Tubes were centrifuged at 1,900 x g for 15 minutes, and the supernatant was transferred and centrifuged again at 1,900 x g for 10 minutes. Plasma was aliquoted and stored at -80 °C. cfDNA was extracted from 3 mL of plasma using the QIASymphony Circulating Nucleic Acid Kit (QIAGEN, Germany). Quantification was performed with the Cell-free DNA ScreenTape Analysis assay on the Agilent 4200 TapeStation System (Agilent Technologies, USA). Libraries were constructed using a minimum of 3 ng cfDNA and the ThruPLEX Plasma-Seq Kit (Takara Bio, Japan), quantified using the D1000/D5000 ScreenTape Analysis assay (Agilent Technologies, USA), and pooled in equimolar concentrations for sequencing.

#### Sequencing and Feature Extraction

WGS was performed on an Illumina NovaSeq 6000/NovaSeq X system using the S4 flow cell (150 bp paired-end reads). Across all LBCL samples, the median sequencing coverage was 1.34X. Quality control was performed using FastQC (v0.11.9), and adapter trimming was performed with BBDuk (BBmap v38.79) using the parameters ktrim=r, k=23, mink=11, hdist=1, and a predefined adapter reference file. Trimmed reads were aligned to the human reference genome GRCh38/hg38 using BWA-MEM (v0.7.17), followed by coordinate sorting with samtools. PCR duplicates were marked using sambamba markdup (v0.8.1), and BAM files were indexed with samtools index (v1.12). To generate high-confidence fragmentomic profiles, aligned reads were additionally filtered using samtools to retain reads with mapping quality MAPQ ≥ 30 and to exclude unmapped, secondary, supplementary, duplicate, and QC-failed reads. To minimize potential confounding effects of sequencing depth, coverage was harmonized across samples.

Mean genome-wide coverage was estimated using samtools depth across all genomic positions, and samples with mean depth above 1x were downsampled to a target mean coverage of 1x using Picard DownsampleSam with the HighAccuracy strategy, where the downsampling probability was defined as:

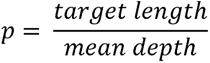

Samples with mean coverage at or below 1x were left unchanged. This ensured comparable sequencing depth across samples for downstream fragmentation analyses.

#### ichorCNA

Tumor fraction estimation was performed using ichorCNA^26^ (v0.3.2-2). First, genome-wide read counts were generated from filtered BAM files using readCounter with a bin size of 1 Mb bins. The resulting coverage profiles were then analyzed with the ichorCNA tool using the hg38/GRCh38 reference genome and an in-house panel of normals derived from 18 healthy control samples. ichorCNA was run with an initial normal fraction of 0.5, initial ploidy of 2, and a maximum copy number of 5. Homozygous deletion calling was disabled, while estimation of normal fraction and ploidy was enabled and estimation of subclonal prevalence was disabled. Chromosomes chr1–chr22 were used for model training. Segmentation parameters were set to txnE=0.9999 and txnStrength=10000, and the male Y-chromosome fraction parameter was set to 0.002. A 3% limit of detection threshold was applied: TF values <3% were classified as negative, whereas TF values ≥3% were classified as positive.

#### Fragment size analysis

Fragment sizes were extracted from paired-end filtered BAM files using the insert size of read pairs, and only fragments between 0 and 400 bp were retained for downstream analysis. For each sample, fragment size distributions were summarized in 20 bp bins across the 0–400 bp range. Per-bin densities were calculated by dividing the number of fragments in each bin by the total number of fragments and by the bin width. These 20 bp bin fractions were compared between non-progressors and progressors in the training set using a two-sided Wilcoxon rank sum test. Three bins (100–120 bp, 120–140 bp, and 180–200 bp) showing significant differences in the training set were selected as fragment size features for downstream modeling.

#### Fragment-end motif analysis

Fragment end sequence analysis was performed using the FrEIA toolkit.^30^ As part of this workflow, for each sample, the first three mapped bases at the 5′ ends of quality-filtered reads were extracted and used to assign each cfDNA fragment a trinucleotide signature. Analyses included fragments with lengths between 1 and 500 bp, without bootstrap resampling. The FrEIA toolkit then generated per-sample motif abundance tables together with downstream summary metrics, including motif diversity score (MDS), Gini index, and the composite FrEIA score. The FrEIA score was computed using a predefined panel of medians together with predefined sets of control-associated and cancer-associated trinucleotides. No batch correction was applied.

Within the FrEIA toolkit, MDS was derived for each sample from fragment-end trinucleotide frequencies as normalized Shannon entropy

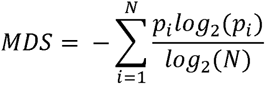

Where *p_i_* denotes the relative abundance of motif *i* among the unique 5′ trinucleotide fragment-end motifs, and *N* is the total number of distinct motifs considered. Higher MDS values indicate a more diverse and even distribution of fragment-end motifs, whereas lower values indicate lower diversity and greater dominance of a subset of motifs.

#### LIONHEART

LIONHEART analysis was performed on filtered BAM files using the authors’ published pipeline^32^ and reference resources to infer the cell and tissue types contributing to cfDNA. The method quantifies genome-wide cfDNA coverage patterns and correlates them with 898 reference chromatin accessibility profiles derived from a broad panel of non-cancerous and cancerous cell lines and tissue types. For each sample, the pipeline produced a feature table in which each column represented a reference cell or tissue type and each value represented the corresponding LIONHEART score. LIONHEART is based on the principle that open-chromatin regions in a contributing cell or tissue type tend to have reduced cfDNA coverage, as these regions are relatively nucleosome-depleted and less protected from fragmentation. Based on this, a more negative LIONHEART score indicates that the sample’s cfDNA coverage pattern is more consistent with contribution from the corresponding cell or tissue type, whereas a more positive score indicates weaker representation of that profile. These scores were used as high-dimensional input features for downstream feature selection and model development.

#### DELFI-FTK

To derive genome-wide regional fragmentation features from cfDNA, DELFI-FTK analysis was performed using the delfi function in FinaleToolkit (v0.11.0).^34^ DELFI-FTK provides an executable implementation of the original DELFI^31^ approach. Briefly, fragmentation features were computed from autosomal genomic bins defined in a 100 kb bins file and merged using the default 5 Mb window size. For each resulting region, the GC-corrected ratio of short to long cfDNA fragments was calculated to generate a regional fragmentation profile for each sample. Analyses were performed on filtered BAM files using the GRCh38 reference together with chromosome sizes, blacklist, and gap annotations. The resulting 511 regional DELFI-FTK features were used as high-dimensional input for downstream feature selection and model development.

#### Feature selection for high-dimensional fragmentomic features

For high-dimensional feature sets, including LIONHEART and DELFI-FTK, feature selection was performed within the training cohort using elastic-net regularized logistic regression. Features were standardized by z-score transformation before model fitting, and hyperparameters controlling regularization strength and the balance between L1 and L2 penalties were optimized by stratified 5-fold cross-validation using ROC AUC as the objective function. Features with non-zero coefficients in the fitted elastic-net model were considered selected. To assess robustness, the stratified cross-validation procedure was repeated ten times using different random shuffles of the training data. Feature selection stability was then summarized by the frequency with which each feature was selected across runs.

For LIONHEART, only seven features had non-zero coefficients and were therefore retained. For DELFI-FTK, features with non-zero coefficients that were retained in at least five of the ten iterations were selected for downstream analysis. The full lists of selected LIONHEART and DELFI-FTK features are provided in **Table S2** and **Table S3**.

#### Multi-Modal cfDNA Analysis and Machine Learning Framework for Outcome Prediction

We developed a machine learning (ML) framework integrating multiple cfDNA features to predict clinical outcomes using the training cohort (Cohort A) and validating it on the independent validation cohort (Cohort B). Specifically, we designed an ML framework to predict 2-year time-to-progression (TTP), using the 6 cfDNA feature sets within a binary classification paradigm. The model outputs a probability score termed the ACT score (Aberrations, Composition of fragments, Terminal motif analyses), derived from a single WGS run of plasma samples collected after one cycle of immunochemotherapy. Samples with ACT scores above a classification threshold are predicted to experience disease progression, while those below the threshold are predicted to be progression-free.

We evaluated six distinct supervised classifiers using a similar framework: Logistic Regression, Random Forest, Gaussian Naive Bayes, Support Vector Machines, Easy Ensemble and Gradient Boosting. Logistic regression demonstrated the best overall performance and was selected as the primary prediction model. Detailed performance metrics for all classifiers are reported in **Table S6**.

Model hyperparameters were optimized using stratified 5-fold cross-validation to maximize prediction accuracy, with the search space defined in **Table S4**. To ensure robustness, we perturbed the training data with ten different random seeds and repeated the optimization process for each, yielding ten alternative optimal models. The classification threshold was also optimized during training and averaged across the ten models. For the independent validation cohort (Cohort B), we averaged the ACT scores from the ten models and applied the corresponding averaged classification threshold to classify samples.

Coefficient extraction was performed only using the training cohort. For each of the ten optimized logistic regression models generated using different random seeds, feature coefficients were extracted from the fitted classifier and then averaged across runs. The sign of each coefficient was used to indicate directionality, with positive coefficients corresponding to features associated with increased predicted risk of progression and negative coefficients corresponding to features associated with decreased predicted risk of progression. Because features were not standardized before model fitting, coefficients were reported on the original feature scale. To aid comparison across features with different scales, we additionally calculated, for each feature, the absolute product of its averaged coefficient and its standard deviation in the training cohort. Averaged coefficients, feature standard deviations, coefficient × standard deviation values, and coefficient directions are reported in **Table S5**.

The ML framework was implemented in Python (v3.12.2) using scikit-learn (v1.5.0), NumPy (v1.26.4), SciPy (v1.11.4), and pandas (v2.2.1).

### Statistical analysis

All statistical analyses and visualizations were performed in R (v4.5.2; R Core Team, Vienna, Austria). All P-values are two-sided. Survival analyses were conducted using the survival (v3.8.6) and survminer (v0.5.1) packages with default settings, employing Kaplan–Meier estimation and Cox proportional hazards modelling. Hazard ratios (HR) with 95% confidence intervals (CI) and log-rank P-values are reported throughout. For boxplots, significance thresholds are denoted as follows: ns, p ≥ 0.05; *, p < 0.05; **, p < 0.01; ***, p < 0.001; ****, p < 0.0001.

## Additional resources

HOVON-902 is registered with number NCT04139252 on ClinicalTrials.gov

HOVON-152 is registered with number NCT03620578 on ClinicalTrials.gov

## Supplemental Information (2)

Document S1. Article plus Supplemental Figures (S1-S6).

Document S2. Supplemental Tables S1–S8.

