## Supplementary files for "Cell-Free DNA Genomic and Fragmentomic Features for Early Outcome Prediction in Large B-Cell Lymphoma"

**Supplemental Information**

**Supplementary Figures**

Figure S1. Geographic distribution of samples across training and validation cohorts.

Figure S2. Baseline fragmentomic profiles in LBCL patients and healthy donors.

Figure S3. Baseline fragmentomic features in LBCL patients by clinical outcome.

Figure S4. Correlation of cfDNA-derived fragmentomic features at T1.

Figure S5. Prognostic performance of IPI and interim PET for risk stratification.

Figure S6. Prognostic performance of ACT score in combination with iPET in Cohort B.

**Supplementary Figures**

**Figure S1. Geographic distribution of samples across training and validation cohorts.**

(A) Cohort A (training) comprises 115 samples collected across 27 hospitals. (B) Cohort B (validation) comprises 59 samples collected across 15 hospitals.

**Figure S2. Baseline fragmentomic profiles in LBCL patients and healthy donors.**

(A) Fragment size distributions in DLBCL patients and HDs, binned at 20 bp intervals. (B) Top ten most differentially expressed LIONHEART features. (C) DELFI-FTK regional cell-free DNA (cfDNA) fragmentation profiles across chromosome arms in DLBCL patients and HDs, displayed as per-sample mean-centered DELFI scores. (D) Principal component analysis (PCA) of DELFI-FTK features.

**Figure S3. Baseline fragmentomic features in LBCL patients by clinical outcome.**

(A) Fragment size distributions in DLBCL patients stratified by clinical outcome (NP, non-progressors vs. P, progressors), binned at 20 bp intervals. (B) Top ten most differentially expressed LIONHEART features. (C) DELFI-FTK regional cfDNA fragmentation profiles across chromosome arms in DLBCL patients stratified by clinical outcome, displayed as per-sample mean-centered DELFI scores. (D) PCA of DELFI-FTK features.

**Figure S4. Correlation of cfDNA-derived fragmentomic features at T1.**

Correlation matrix of six cfDNA-derived features measured at T1 in the Training cohort (n = 115): TF (tumor fraction by ichorCNA), three fragment-size bins significantly different between non-progressors and progressors (100–120, 120–140, 180–200 bp), MDS (motif diversity score), and FrEIA (4-mer end-motif bias score). Cell color and value indicate Spearman's ρ; stars denote significance (* p < 0.05, ** p < 0.01, *** p < 0.001).

**Figure S5. Prognostic performance of IPI and interim PET for risk stratification.**

(A) Kaplan–Meier estimates of time to progression (TTP) stratified by International Prognostic Index (IPI) (0-2 vs. 3-5) in all patients with available IPI at T1 (n=173). (B) Kaplan–Meier estimates of overall survival (OS) stratified by IPI (0-2 vs. 3-5) in all patients with available IPI at T1 (n=173). (C) Kaplan–Meier estimates of TTP stratified by interim PET-CT (iPET) results (CMR vs non-CMR) in HGBL patients (n=87). (D) Kaplan–Meier estimates of OS stratified by iPET results (CMR vs non-CMR) in HGBL patients (n=87). CMR corresponds to a Deauville score of 1–3; non-CMR, corresponds to a Deauville score of 4–5. iPET was performed after three cycles of immunochemotherapy and was limited to high-grade B-cell lymphoma (HGBL) patients enrolled in the HOVON-152 trial; Hazard ratio (HR) with 95% confidence interval (CI) and log-rank P-value are indicated.

**Figure S6. Prognostic performance of ACT score in combination with iPET in Cohort B.**

(A) Kaplan–Meier estimates of time-to-progression (TTP) stratified by iPET in patients with both IPI and ACT score available in Cohort B (n=32). (B) Kaplan–Meier estimates of overall survival (OS) stratified by iPET in patients with both iPET and ACT score available in Cohort B (n=32). (C) Kaplan–Meier estimates of TTP stratified by combined ACT score and iPET in patients with both metrics available in Cohort B (n=32). (D) Kaplan–Meier estimates of OS stratified by combined ACT score and iPET in patients with both metrics available in Cohort B (n=32). (E) Forest plot of multivariable Cox proportional hazards analysis for TTP in Cohort B, including iPET+ and ACT+ as covariates. Negative iPET is defined as complete metabolic response (CMR) (Deauville 0-3); Positive iPET is defined as non-CMR (Deauville 4-5). Hazard ratio (HR) with 95% confidence interval (CI) and log-rank *P*-value are indicated for all Kaplan–Meier analyses.
