## Supplementary figures and images for "Cell-Free DNA Genomic and Fragmentomic Features for Early Outcome Prediction in Large B-Cell Lymphoma"

### Figure S1

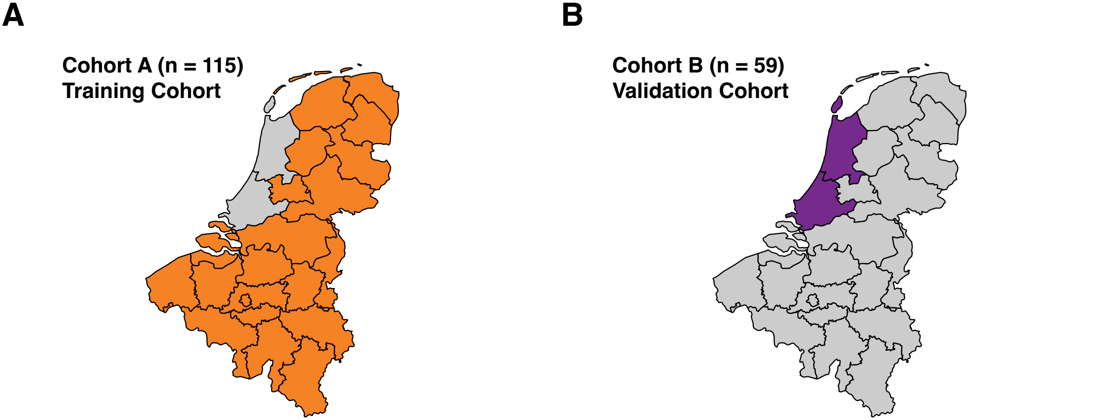

### Figure S2

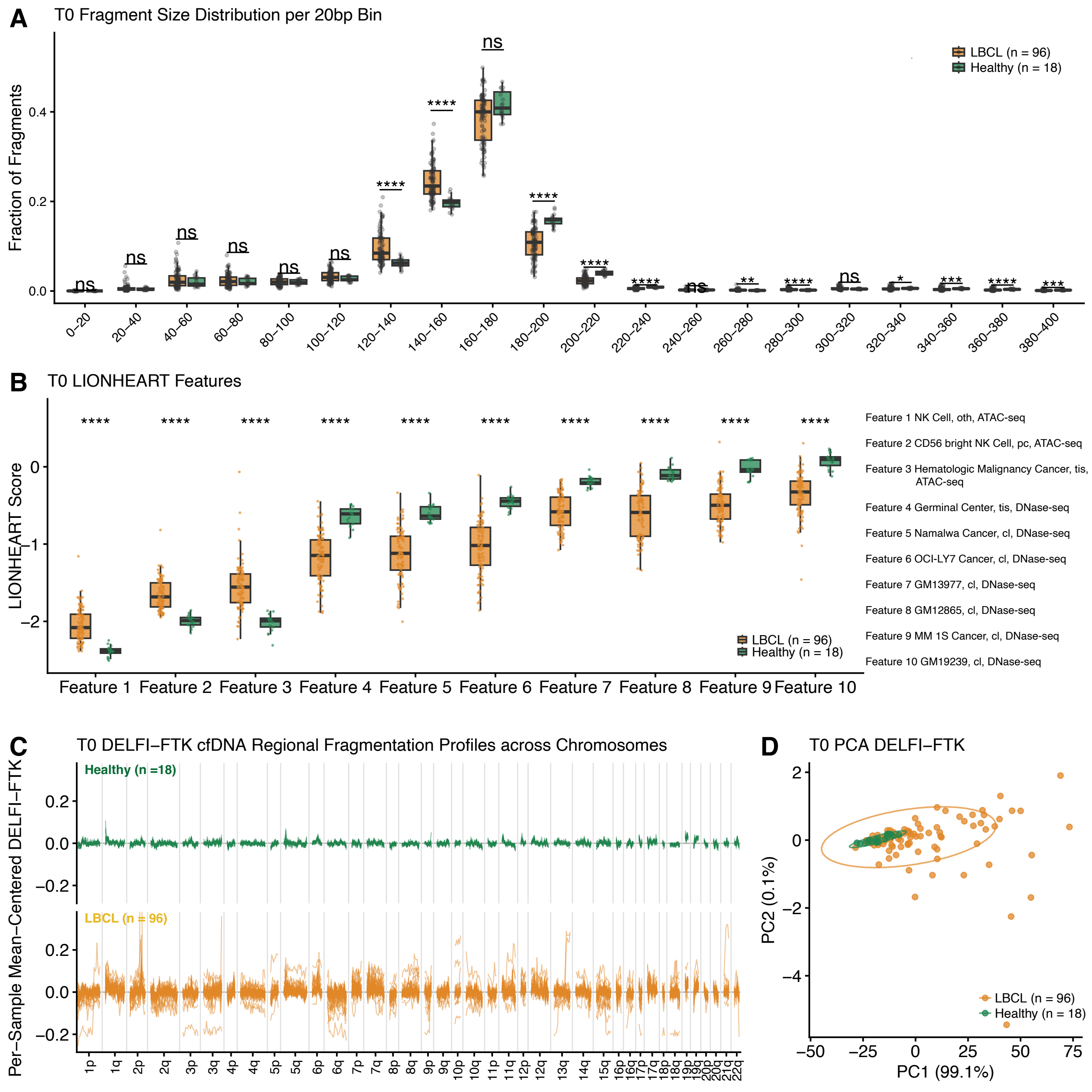

### Figure S3

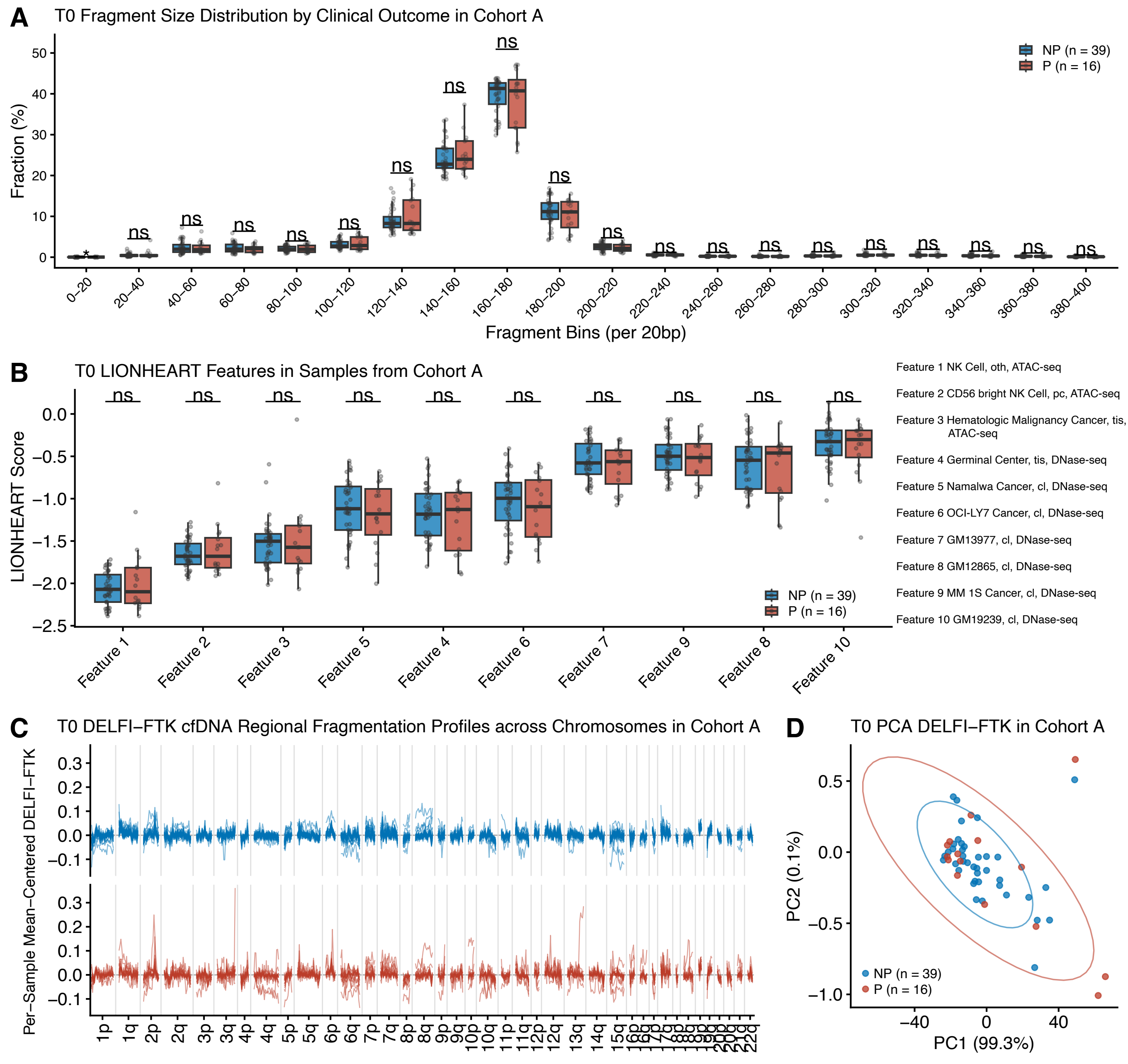

### Figure S4

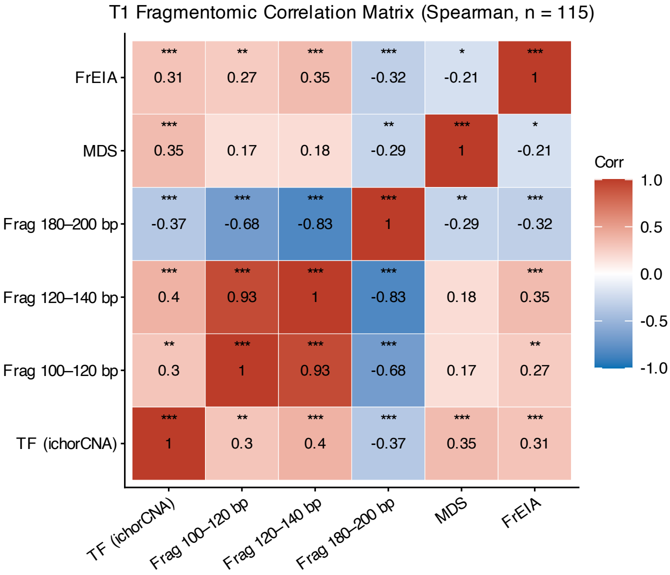

### Figure S5

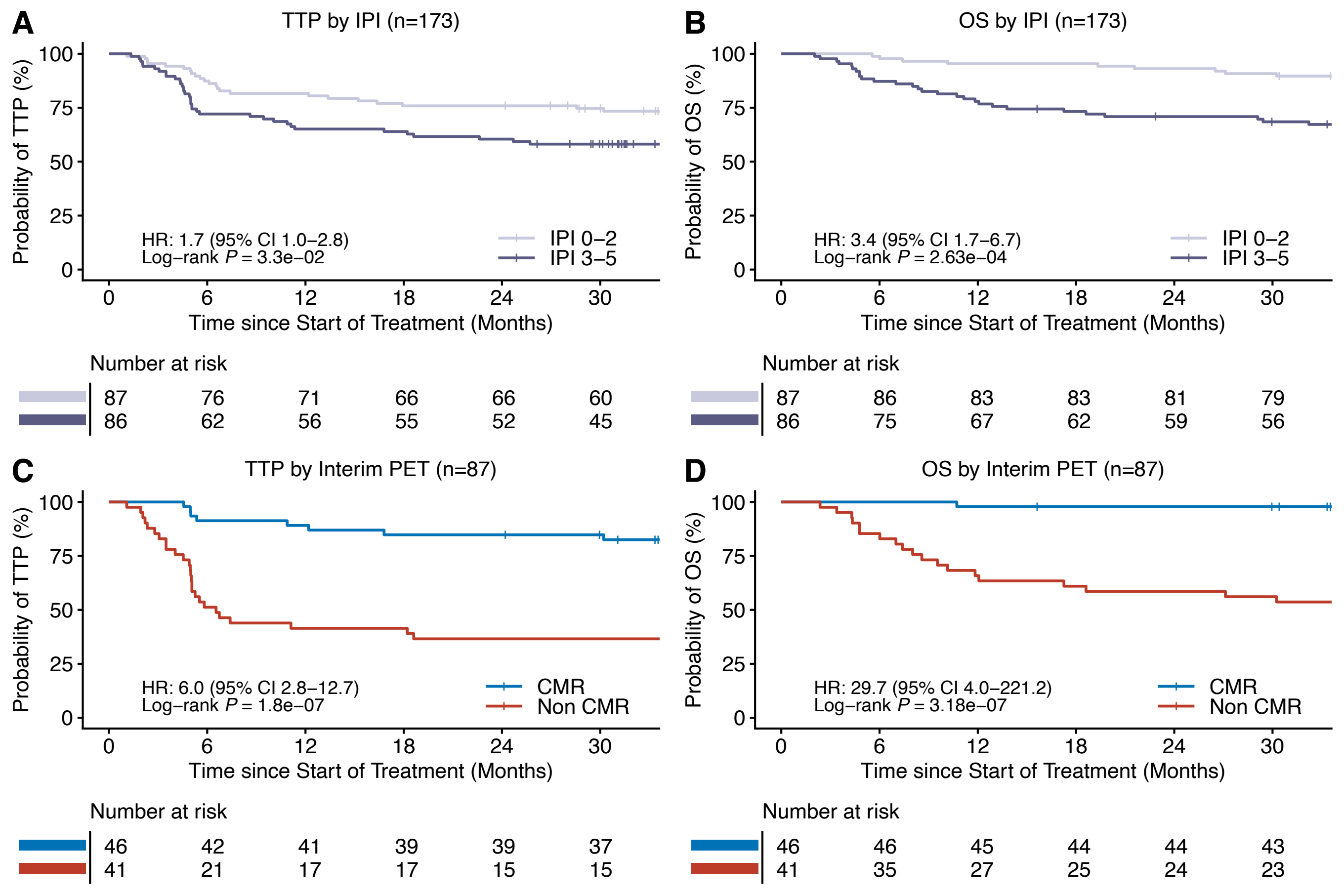

### Figure S6

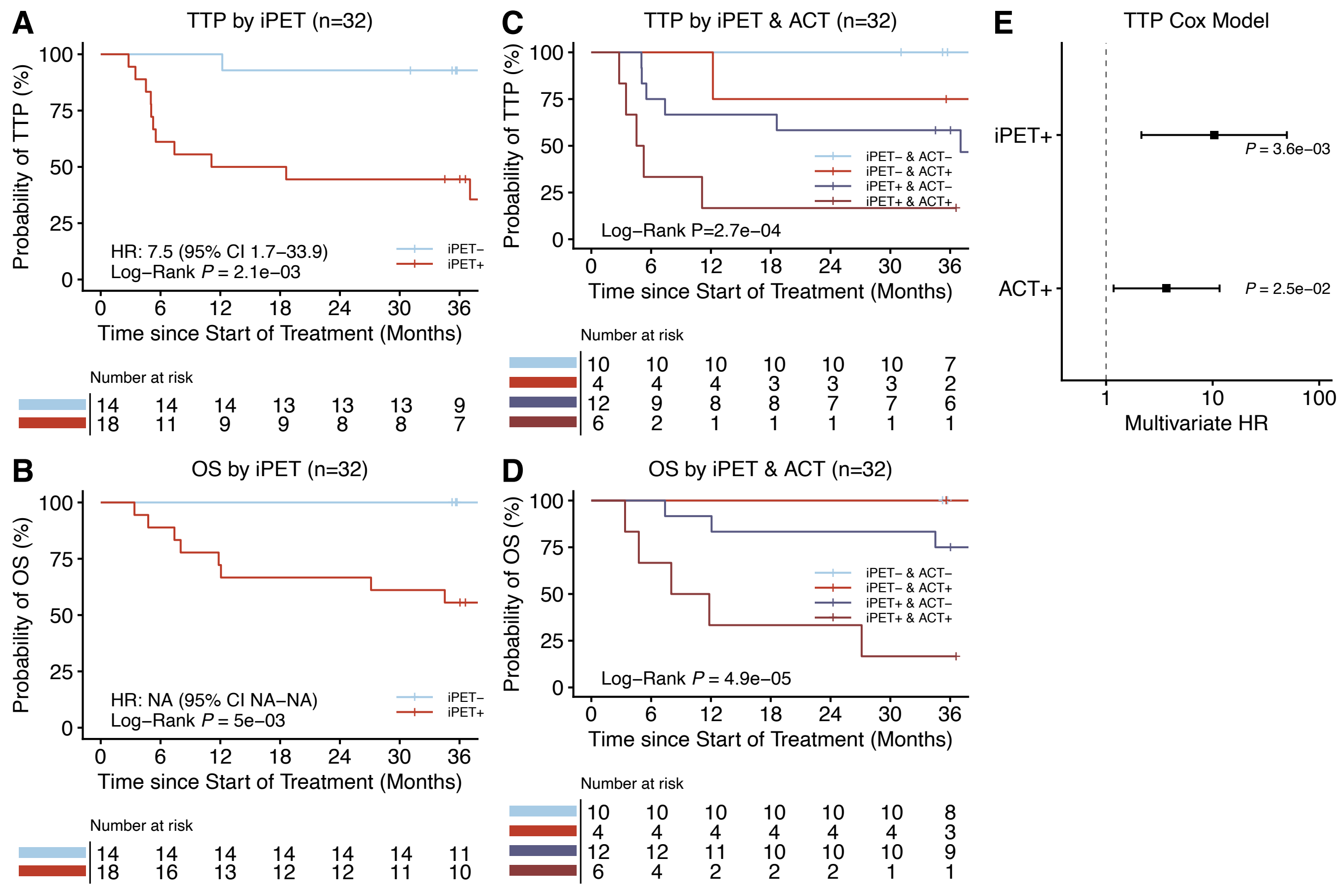
